# Effects of bilateral sequential theta-burst stimulation on 5-HT_1A_ receptors on dorsolateral prefrontal cortex in treatment resistant depression

**DOI:** 10.1101/2022.02.18.22271165

**Authors:** Matej Murgaš, Jakob Unterholzner, Peter Stöhrmann, Cécile Philippe, Godber M. Godbersen, Lukas Nics, Murray B. Reed, Chrysoula Vraka, Thomas Vanicek, Wolfgang Wadsak, Georg S. Kranz, Andreas Hahn, Markus Mitterhauser, Marcus Hacker, Siegfried Kasper, Rupert Lanzenberger, Pia Baldinger-Melich

**Affiliations:** Department of Psychiatry and Psychotherapy, Clinical Division of General Psychiatry, Medical University of Vienna, Austria; Department of Biomedical Imaging and Image-guided Therapy, Division of Nuclear Medicine, Medical University of Vienna, Austria; Ludwig Boltzmann Institute Applied Diagnostics, Vienna, Austria; Department of Rehabilitation Sciences, The Hong Kong Polytechnic University, Hung Hom, Hong Kong; Department of Chemistry, Institute of Inorganic Chemistry, University of Vienna, Austria

**Keywords:** Theta-burst stimulation, Transcranial magnetic stimulation, Treatment-resistant depression, Serotonin-1A receptor, Positron emission tomography

## Abstract

Theta-burst stimulation (TBS) represents a brain stimulation technique effective for treatment-resistant depression (TRD) as underlined by meta-analyses. While the methodology undergoes constant refinement, bilateral stimulation of the dorsolateral prefrontal cortex (DLPFC) appears promising to restore left DLPFC hypoactivity and right hyperactivity found in depression. The post-synaptic inhibitory serotonin-1A (5-HT_1A_) receptor, also occurring in the DLPFC, might be involved in this mechanism of action. To test this hypothesis, we performed PET-imaging using the tracer [*carbonyl*-^11^C]WAY-100635 including arterial blood sampling before and after a three-week treatment with TBS in 11 TRD patients compared to sham stimulation (n=8 and n=3, respectively). Treatment groups were randomly assigned, and TBS protocol consisted in excitatory intermittent TBS to the left and inhibitory continuous TBS to the right DLPFC. A linear mixed model including group, hemisphere time and Hamilton Rating Scale for Depression (HAMD) score revealed a 3-way interaction effect of group time and HAMD on 5-HT_1A_ receptor specific binding V_S_. While post-hoc comparisons showed no significant changes of 5-HT_1A_ V_S_ in either group, higher 5-HT_1A_ V_S_ after treatment correlated with greater difference in HAMD (r=-0.62), indicative of potential effects of TBS on the 5-HT_1A_ receptor. Due to the small sample size, all results, however, must be regarded with caution.

## INTRODUCTION

Major depressive disorder (MDD) is a common, debilitating psychiatric illness that – along with personal suffering and psychosocial strain – represents an immense socioeconomic burden worldwide [1, 2]. MDD is a treatable disorder with pharmacological and psychotherapeutic interventions constituting the fundamental pillars of antidepressant treatment. Nevertheless, up to 60% of the patients do not satisfactorily respond to first-line pharmacological treatments [3]: This subgroup of patients is conventionally labeled as treatment-resistant, which implies the failure of response to at least two adequate antidepressant trials [4]. The significant amount of affected persons calls for an improvement of treatment outcomes by exploring alternative treatment strategies, such as novel rapid-acting antidepressants like esketamine and improved brain stimulation techniques [5-7].

Theta-burst stimulation (TBS), an enhanced derivative of repetitive transcranial magnetic stimulation (rTMS), is an effective non-pharmacological treatment option for MDD, combining the approved efficacy of rTMS while offering better practicability with significantly shorter therapy duration [8, 9]. Several double-blind, sham-controlled, multicenter trials as well as meta-analyses have provided evidence for the antidepressant effects of rTMS [10, 11], which has been approved by the United States Food and Drug Administration (FDA) as a therapeutic option for treatment-resistant depression (TRD) since 2008 [12, 13]. Since the development of this noninvasive brain stimulation technique in the 1980s [14], research into therapeutic TMS for a multitude of neurological and psychiatric disorders, particularly depression [15], has dramatically increased; its principle lies in the induction of electrical currents inside the brain by electromagnetic pulses applied on the scalp, causing neuronal depolarization and functional alteration of brain activity in specifically targeted regions.

A key variable in TMS is the application frequency of electromagnetic pulses. While low frequency or single pulses were shown to decrease brain activity in the stimulated region [16], high frequency pulses exhibit excitatory properties e.g., the FDA-approved 10-Hz or high-frequency (HF) TMS over the left dorsolateral prefrontal cortex (DLFPC) in depression [17]. Similarly, two protocols of TBS with opposing effects on cortical excitability have been proposed; intermittent (iTBS) and continuous TBS (cTBS) with excitatory and inhibitory consequences, respectively. Bilateral TBS, which combines iTBS to the left, alleged hypoactive DLPFC and cTBS to the right, alleged hyperactive DLPFC holds promise to be the most efficacious neuromodulation measure in TRD, thereby even challenging the panacea electroconvulsive therapy (ECT) [18, 19].

Though the exact neurobiological mechanisms by which TMS alters mood remain to be elucidated, the commonly acknowledged explanation involves therapeutic neuroplasticity through long-lasting modulation of cortical excitability that go beyond the stimulated brain region [20-22]. To investigate the TMS-induced changes in neural activity patterns, several neuroimaging studies have been performed to identify brain networks involved in its antidepressant effects and inform personalized approaches in the future [23-26]. The vast majority of the trials using positron emission tomography (PET) focused on measures of cerebral blood flow [27-30] and cerebral glucose metabolism [31], showing changes in neural network dynamics beyond the cortical site directly targeted by the electromagnetic pulse. Regarding effects on modulatory neurotransmitter systems, rTMS was shown to be accompanied by increases in extracellular dopamine in the stimulated hemisphere (basal ganglia, anterior cingulate and medial orbitofrontal cortex) measured using [^11^C]raclopride [32-34], significant changes in regional serotonin synthesis capacity in limbic areas assessed using alpha-[^11^C]-methyltryptophan [35] as well as a serum serotonin level enhancement [36]. Preclinical investigations in rats indicate that TMS might exert its antidepressant effects via modulation of the serotonergic system [37, 38], particularly the inhibitory serotonin-1A (5-HT_1A_) [38-41]. This receptor is prone to profound changes in mood and anxiety disorders [42-45] and represents an important player of antidepressant pharmacotherapy and electroconvulsive therapy in humans as shown previously by our group [46-49]. Based on this evidence, we aimed at assessing the impact of bilateral TBS on 5-HT_1A_ receptor distribution volume in a sample of TRD patients using the radioligand [*carbonyl*-^11^C]WAY-100635 to probe the hypothesis of a 5-HT_1A_ receptor reduction in the DLFPC – as seen with other antidepressant treatments – by this non-invasive brain stimulation technique *in vivo*.

## MATERIAL and METHODS

### Subjects and study design

35 subjects suffering from treatment-resistant depression (defined as failure to respond for the current episode to two adequate medication trials of at least 4 weeks in sufficient dosage) were recruited via the outpatient department and the hospital wards of the Department of Psychiatry and Psychotherapy at the Medical University of Vienna, Austria, and enrolled in the study (ClinicalTrials.gov Identifier NCT02810717). 24 subjects dropped out of the study, mainly due to technical and schedule planning issues, leaving a final sample size of 11. In this randomized and double-blind clinical trial patients received either bilateral Theta-burst (TBS, n=8) or sham (n=3) stimulation. Each participant underwent PET measurement with [*carbonyl*-^11^C]WAY-100635 once before (PET1) and once after TBS treatment (PET2). In addition, structural images were recorded using magnetic resonance imaging (MRI) scans at each PET scanning session, which were used for Neuronavigation and co-registration of dynamic PET data.

Subjects were carefully screened by a psychiatrist and included in the trial when fulfilling criteria for a single or recurrent major depression (using the Structural Clinical Interview for DSM IV Diagnosis, SCID IV) and a 17-item Hamilton Rating Scale for Depression (HAMD) score ≥ 18 (at least moderate depression) assessed at the inclusion as well as on the individual measurement days. Concomitant antidepressant treatment was allowed, if stable, four weeks prior study enrolment und during study participation. Exclusion criteria were major systemic (untreated) or neurological disorders, including brain injuries, current substance abuse (ruled out using SCID IV and a urine drug screening), current psychotic symptoms, pregnancy and any contraindication for magnetic resonance imaging or TMS [50]. Also, intake within four weeks prior the first examination visit or current intake of psychotropic drugs targeting the 5-HT_1A_ receptor (i.e. clozapine, aripiprazole, quetiapine (>100 mg), ziprasidone, amitriptyline, nebivolol, propranolol, mirtazapine, triptans, trazodone) was considered as an exclusion criterion. A causal relationship of mood disturbances and general medical conditions was further ruled out by clinical examination, routine laboratory measurements (complete blood cell count, chemistry, thyroid hormones) and an electrocardiogram.

Study data were collected and managed using REDCap electronic data capture tools hosted at the Department of Psychiatry and Psychotherapy, Medical University of Vienna, Austria [51, 52]. The study was approved by the ethics committee of the Medical University of Vienna, Austria (1761/2015). Each subject provided written informed consent and was financially reimbursed for the participation in the study. Procedures performed during the study were in accordance with the Declaration of Helsinki including all revisions.

### Theta-burst stimulation treatment

Patients were treated with bilateral TBS or sham TBS using MagPro X100 model (MagVenture, Tonica Elektronik A/S, Denmark, www.tonika.dk) and a Cool-B70 Butterfly coil. For each treatment session intermittent TBS (iTBS) was applied to the left dorsolateral prefrontal cortex (DLPFC), whereas continuous TBS (cTBS) was applied to the right DLPFC at an intensity of 120% resting motor threshold for the first dorsal interosseous muscle [53]. The iTBS consisted of 2-second trains with an inter-train-interval of 8 seconds. Trains (30 pulses, 10 bursts) were repeated 20 times to reach a total number of 600 pulses per session. The cTBS comprised uninterrupted bursts reaching a total number of 600 pulses per session. For both iTBS and cTBS 3-pulse 50-Hz bursts were given every 200 ms [9]. Two sessions, each lasting ∼5 minutes, were scheduled daily, given 60 minutes apart [54], Monday to Friday, for three weeks resulting in a minimum of 30 sessions per subjects. Coil placement to the left and right DLPFC was performed using neuro-navigation (Brainsight, LOCALITE® TMS Navigator, Germany [55]) based on MNI coordinates x=±38, y=44, z=26 from individual MR images. Sham TBS comprised bursts as given above with the coil set at 90° against the skull. Thus, sham stimulation was accompanied by similar auditory (clicking noise) and somatosensory (i.e. pricking) artefacts. Patients were blind to the individual group assignment. Efficacy outcome measures were assessed by blinded raters, who were not permitted access to the treatment sessions.

### Neuroimaging

Each PET scan was conducted using a GE Advance PET scanner (General Electric Medical Systems, Milwaukee, Wisconsin) at the Department of Biomedical Imaging and Image-guided Therapy, Division of Nuclear Medicine, Medical University of Vienna, Austria as previously described [48, 49, 56, 57]. To correct for tissue attenuation, 5-minute transmission scan was carried in 2-D mode (retractable 68Ge rod sources). Afterwards, PET measurement started with the bolus administration of [*carbonyl*-^11^C]WAY-100635 (injection dose 4.6 MBq/kg body weight) in cubital vein. All scans were acquired in 3-D mode for 90 minutes (51 frames: 12×5 s, 6×10 s, 3×20 s, 6×30 s, 9×60 s, 15×300 s) and were reconstructed (iterative filtered back-projection algorithm) to final images comprising a spatial resolution of 4.36mm full-width at half-maximum 1 cm next to the center of the field of view (matrix 128×128, 35 slices). The radioligand [*carbonyl*-11C]WAY-100635 was prepared according to previously published methods [58] at the Cyclotron Unit of the PET Center. Each PET scan was complemented with arterial blood samples for the quantification of [*carbonyl*-^11^C]WAY-100635 that were automatically drawn for first 10 minutes (ALLOGG, Mariefred, Sweden) and manually at 2, 5, 6, 7, 8, 10, 20, 40 and 60 minute of the measurement.

Structural T1 weighted MR image were acquired at both PET measurements with the magnetization prepared rapid gradient echo sequence (MP-RAGE with TE/TR = 4.21/3000 ms, voxel size 1 × 1 × 1.1 mm3) using a 3 T PRISMA MR Scanner (Siemens Medical, Erlangen, Germany).

### Data processing and quantification

Following correction for tissue attenuation, PET scan of each patient was corrected for head motion, co-registered to the structural T1-weighted image. The latter was afterwards normalized to MNI space producing a transformation matrix that was further applied to normalize co-registered PET data to MNI space. All preprocessing steps were done using SPM (Wellcome Trust Centre for Neuroimaging, London, United Kingdom; http://www.fil.ion.ucl.ac.uk/spm/) and Matlab 2018a (The Mathworks Inc., Natick, MA, USA). Subsequently, time activity curves (TACs) were extracted for selected regions of interest (ROIs) - left and right DLPFC and cerebellar white matter (CWM). DLPFC ROIs were defined as a sphere with diameter of 10 mm around the MNI coordinate representing the individual application point of TBS treatment. The CWM ROI was extracted using an in-house created atlas [59]. To reduce the noise induced by short frames in the beginning of the scan, the first 2 minutes (frames 12×5 s and 6×10 s) of the measurement were resampled to 20-second frames.

The arterial input functions representing non-metabolized radioligand in plasma were obtained as product of the whole blood activity, plasma-to-whole blood ratio (average) and fraction of intact radioligand in the plasma (fitted with the Hill-type function). Afterwards, the distribution volume of specifically bound ligand V_S_, representing the distribution of 5-HT_1A_ receptor, was estimated for DLPFC. Quantification of 5-HT_1A_ V_S_ was carried out utilizing a constrained 2 two-tissue compartment model. Here, CWM was fitted and the ratio of K_1_/k_2_ was fixed for the DLPFC regions [60].

### Statistical analysis

All statistical analyses were performed in SPSS version 28 for Windows (SPSS Inc., Chicago, Illinois; www.spss.com). A linear mixed model was used to assess the effect of TBS treatment on 5-HT_1A_ receptor V_S_ in the DLPFC in TRD patients using group assignment (TBS or sham), time point of measurement (PET 1 or PET 2) and hemisphere of ROI as fixed factors and HAMD scores, representing a scale predictor, as covariate. Of note, the factor hemisphere was introduced in the statistical model as iTBS and cTBS to the left and right DLPFC, respectively, are presumed to display opposing effects on brain activation, thereby potentially bearing lateralized effects on 5-HT_1A_ receptor distribution.

The Mann-Whitney U-test was utilized to assess possible difference in the baseline HAMD score between both groups. Post-hoc exploratory tests for the interactions were done using Wilcoxon Signed Ranked Test. The relationship between V_S_ and HAMD was investigated using the Spearman’s Rank correlation via change in HAMD (*ΔHAMD*= *HAMDPET*2 − *HAMD*_*PET*1_) and the change in V_S_ (*ΔV*_*S*_ = *V*_*S*_*PET*2_ − *V*_*S*_*PET*1_) for the verum group.

In addition, lateralization quotient describing the difference between the activation in left and right hemisphere (LQ [61]) was calculated for each time point (PET1 and PET2) for TBS group for DLPFC (see Table 2).

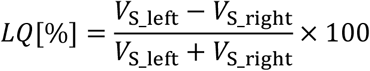

**Table 1.** Demographic information about the patients included in the study.

| Group | TBS | Sham |
| --- | --- | --- |
| n | 8 (5 female) | 3 (3 female) |
| age | $35.88 \pm 8.41$ | $40.33 \pm 6.03$ |
| baseline HAMD (PET 1) | $18.50 \pm 3.55$ | $22.00 \pm 2.65$ |
| HAMD after treatment (PET 2) | $12.12 \pm 5.44$ | $17.33 \pm 10.02$ |
| number of responders | 2 | 1 |

**Table 2.**
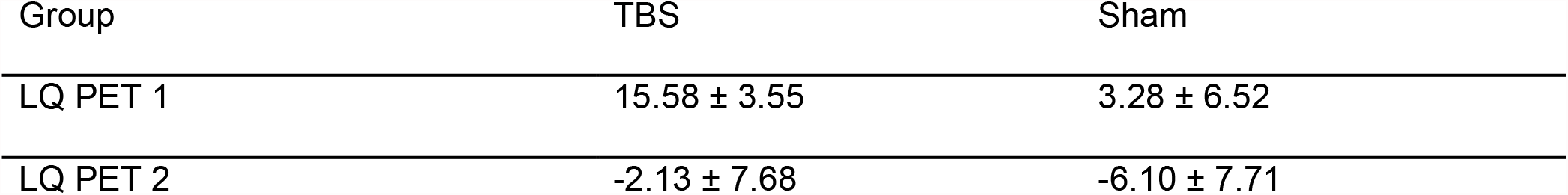
Average lateralization quotient of V_S_ was calculated for each time point and each group separately.

| Group | TBS | Sham |
| --- | --- | --- |
| LQ PET 1 | 15.58 ± 3.55 | 3.28 ± 6.52 |
| LQ PET 2 | -2.13 ± 7.68 | -6.10 ± 7.71 |

Afterwards, a Wilcoxon Signed Ranks Test was used for possible changes in LQ between PET 1 and PET 2. All statistical tests were assessed on the significance level 0.05. No further corrections for multiple testing were done, as the analysis is of exploratory nature.

## RESULTS

Data from eleven TRD subjects were available to examine the impact of three weeks of TBS on 5-HT_1A_ V_S_ in the left and right DLPFC. The sample’s demographics are summarized in Table 1. Eight subjects (five women) aged 35.9 ± 8.4 received i/cTBS, three subjects (only women) aged 40.3 ± 8.0 received sham stimulation. Mean baseline HAMD scores were 18.5 ± 3.6 and 22.0 ± 2.7, respectively, and comparable in both groups (Mann Whitney U test, p=0.18). Concomitant medication of the participants is subsumed in the supplementary Table.

Response to treatment was defined as a reduction of baseline HAMD ≥ 50%. 2 out of 8 (25%) TBS-treated subjects fulfilled these criteria at PET 2 (after TBS), 1 out of 3 (33%) in the sham group.

Linear mixed model analysis using group (TBS vs. sham), time (PET 1 vs. PET 2), hemisphere (left vs. right) and HAMD score showed a main effect of group (F=6.75, p=0.019), time (F=7.45, p=0.015) and HAMD (F=11.00, p=0.004) on 5-HT_1A_ V_S_ as well as two-way interactions between group*time (F=6.24, p=0.024), time*HAMD (F=7.30, p=0.015), group*HAMD (F=6.10, p=0.025), and a three-way interaction between group*time*HAMD (F=6.02, p=0.025). All other two- or three-way interactions and the main effect of hemisphere were non-significant. Post-hoc comparisons using Wilcoxon Signed Ranks test revealed no significant changes of 5-HT_1A_ V_S_ at PET 2 compared to PET 1 in the TBS (p=0.67) and sham group (p=1.00). The estimates of 5-HT_1A_ V_S_ (averaged over hemispheres) were 3.21 ± 1.40 at PET 1 and 3.42 ± 0.80 at PET2 in the TBS group, and 3.13 ± 2.05 at PET 1 and 3.46 ± 0.52 at PET 2 in sham group (see Figure 1).

**Figure 1.**
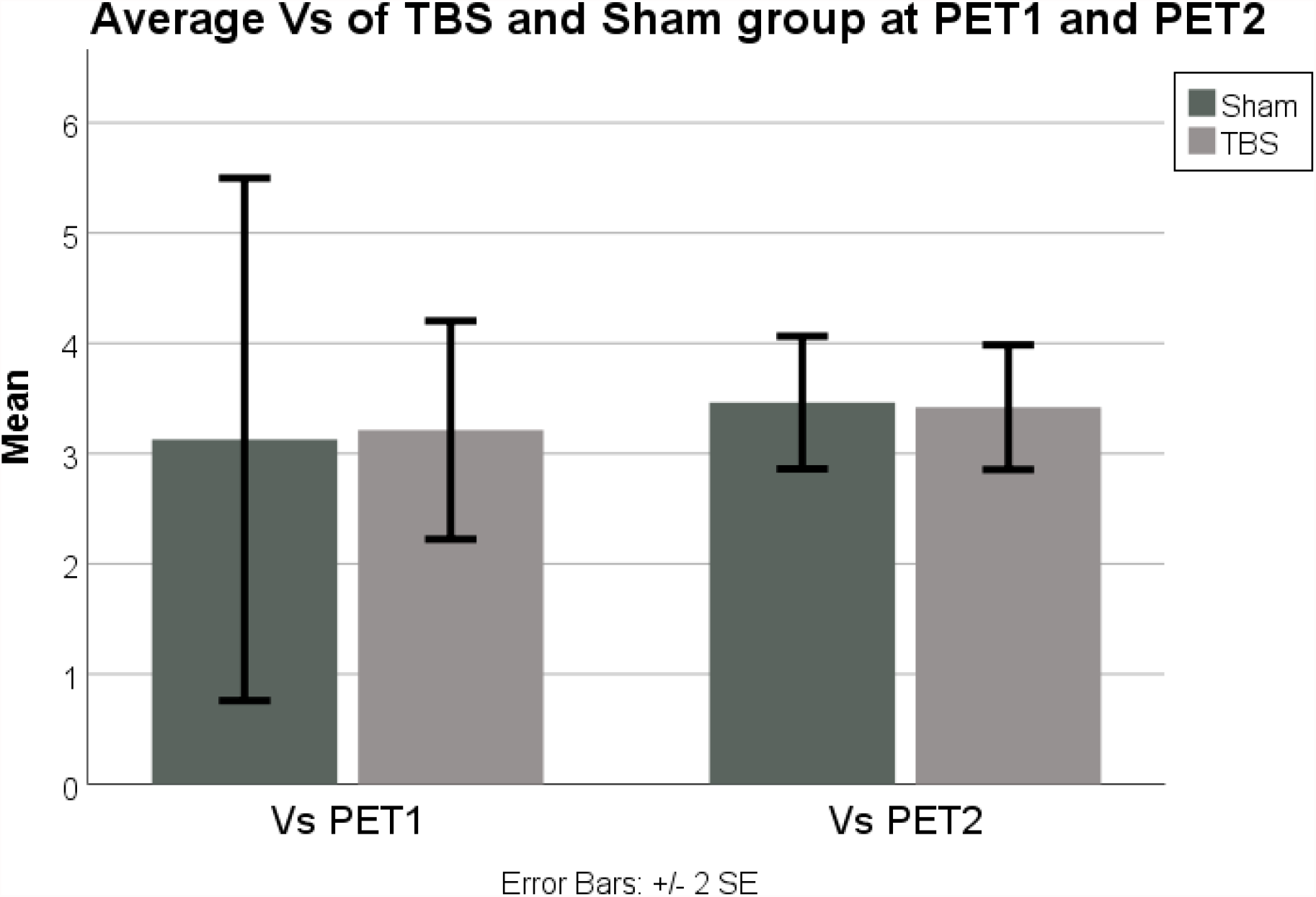
Average 5-HT_1A_ V_S_ in DLPFC for TBS and Sham group at both measurement time points.

Spearman’s rank correlation between change of 5-HT_1A_ V_S_ and ΔHAMD between both PET measurements revealed a negative correlation in the TBS group (r=-0.62, p=0.999) (see Figure 2). Due to the small sample size, this correlation is not reported for the sham group (n=3).

**Figure 2.**
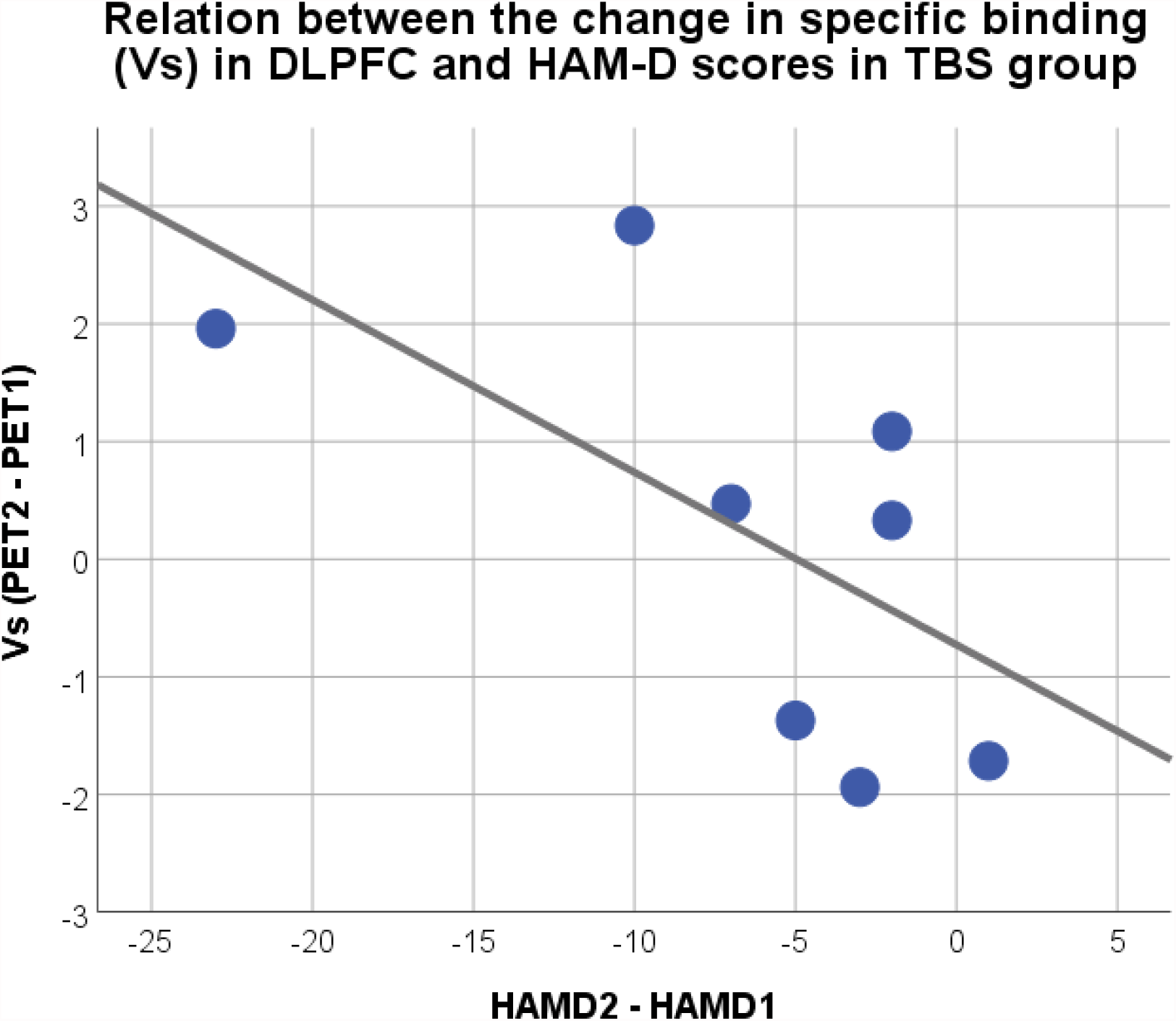
Scatter plot showing the relationship between the ΔHAMD score and ΔV_S_ between PET1 and PET2 in the TBS group. Spearman’s rank correlation showed r=-0.66 (p=0.099).

LQ was computed separately for each group (see Table 2) and did not change following treatment in the TBS group (Wilcoxon Signed Rank test, p=0.069). Due to the small sample size, we did not perform this test in the sham group (n=3).

## DISCUSSION

Specific distribution volumes of 5-HT_1A_ in the stimulation epicenters located in the left and right DLPFC as assessed using [*carbonyl*-^11^C]WAY-100635 appeared to be differentially affected by three weeks of bilateral TBS treatment (iTBS over the left and cTBS over the right DLPFC) compared to sham stimulation in a sample of eleven TRD patients. Given the small sample size particularly in the sham group, the results of this longitudinal PET study should be considered exploratory and must therefore be interpreted carefully.

Based on earlier imaging findings published by our group using the same radioligand (see below), we expected a reduction of 5-HT_1A_ V_S_ in the target region upon completion of TBS. In fact, a reduced availability of 5-HT_1A_ receptors might represent a common neural ground for pharmacological and non-pharmacological antidepressant treatments. Three months of escitalopram intake in patients with anxiety disorders were accompanied by significant reductions in binding potentials in limbic regions [47]. The same direction of change was shown in the raphe in a medication-free depressed sample after SSRI treatment [62]. Interestingly, reductions of 5-HT_1A_ receptor binding were also observed in cortical brain regions following eight weeks of daily intake of Silexan^®^, an anxiolytic herbal compound of lavender essential oil [49]. Most pronounced and widespread cortical reductions of 5-HT_1A_ receptor binding (∼30%) were reported following a course of ECT [48]. However, in none of the studies mentioned above was the degree of binding reductions over time correlated with treatment outcomes.

In the present study, though post-hoc tests did not reveal significant differences in 5-HT_1A_ V_S_ between both PET scans, absolute numbers indicate a slight and unexpected increase of outcome measures in the stimulation epicenters located in the DLPFC in both groups. In addition, the Spearman’s rank correlation between the change of 5-HT_1A_ V_S_ and HAMD after the treatment course suggests the greater the increase of Vs after the treatment course, the greater the reduction of HAMD scores (and the greater the response). Since we did not find significant differences between hemispheres, left iTBS and right cTBS seem to similarly affect the distribution of the 5-HT_1A_ receptor. Given the general assumption that activation of 5-HT_1A_ receptors in projection areas mediates a hyperpolarizing response to serotonin on pyramidal neurons and GABA-ergic neurons [63], an increase of 5-HT_1A_ availability in the DLPFC might result in a disinhibition of neurotransmission and increase in neuronal activity. Though highly speculative, in theory, this constitutes the desired effect of iTBS to the left DLPFC in depression [18]. Currently, no previous *in vivo* data exists to explain our results; however, novel cellular models might be promising to test this hypothesis [64]. Contrasting results regarding 5-HT_1A_ receptor binding might arise through differences in the methodology used, including the choice of reference region and modeling [65, 66].

Still, there is a high level of preclinical evidence supporting our hypothesis of TMS-induced changes within the serotonergic system. Several animal studies have indicated that rTMS may affect the serotonergic system through the 5-HT_1A_ receptors, the most important inhibitory receptor subtype within the serotonergic receptor family, either expressed as an autoreceptor on presynaptic serotonergic neurons or as a heteroreceptor on postsynaptic neurons in projection sites [45]. Single rapid-rate rTMS exposure led to significant increases in 5-HT_1A_ receptors in the frontal cortex as quantified by in-vitro autoradiography 24h after the intervention in rats [38]. Chronic rTMS reduced the ability of OH-DPAT, a full 5-HT_1A_ receptor agonist, to decrease serotonin levels in projection sites, which is indicative of a reduced sensitivity of 5-HT_1A_ autoreceptors [40]. Investigations using intracerebral microdialysis indicate the selective release of monoamines following rTMS, however not necessarily of serotonin [38, 67, 68]. Only few reports show an rTMS-induced serotonin level increase in the rat hippocampus [37, 69] and nucleus accumbens [70], an effect that was also reported in humans [36]. In contrast to the 5-HT_1A_ receptor, the major excitatory serotonin receptor 5-HT_2A_ was shown to be downregulated by chronic rTMS in rats [71]; interestingly, in humans, decrease of 5-HT_2A_ receptors in the hippocampus and the bilateral DLPFC was correlated with treatment response to HF rTMS [72]. Finally, evidence retrieved from genetic investigations emphasizes the association of the 5-HT_1A_ receptor and TMS, as the genotype of the 5-HT_1A_ receptor promoter region polymorphism (rs6295) was shown to influence the outcome of HF TMS in patients suffering from a major depressive episode [73, 74]. It has been suggested that a greater load of G alleles in a 5-HT_1A_ receptor promotor polymorphism might be associated with lower serotonin release, resulting in a post-synaptic upregulation of the 5-HT_1A_ receptor [65]. This polymorphism was, however, not assessed in our study sample.

The effect of TMS in TRD is frequently associated with plastic changes affecting synapse formation, long-term potentiation (LTP) and depression (LTD) [75, 76]. In a study in mice, Cambiaghi et al. have shown, for example, that high frequency rTMS leads to an increase in dendritic complexity in layer II/III pyramidal neurons of the primary motor cortex [77]. The 5-HT_1A_ receptor, in combination with 5-HT signaling, has been repeatedly implicated in plastic changes, including alterations in gray matter volumes [78, 79]. Since the receptor is also located on pyramidal neurons in layers III [80], rTMS might influence its expression and thus mediate its effect on synaptogenesis. Of note, rTMS of glial cells affects neuronal excitability and might, through the presence of 5-HT_1A_ receptors on glial cells [81], lead to alterations in 5-HT_1A_ availability and thus contribute to neuroplasticity [82]. Data on the interplay of rTMS and 5-HT_1A_ receptors in pyramidal and glial cells in cell cultures is however missing up until now [81].

According to the chosen treatment protocol in this study, applying (excitatory) iTBS to the left DLPFC and (inhibiting) cTBS to the right DLPFC, we would have expected a clearer change in LQ between PET 1 and PET 2 in the TBS group. However, the latter was not significant. Considering the absolute values of the computed index, the LQ was positive at baseline in both groups, implying a higher 5-HT_1A_ receptor binding on the left compared to the right hemisphere in symptomatic, depressed patients, and negative following three weeks of treatment, corresponding to a 5-HT_1A_ distribution reversal that seems more pronounced in the TBS group. This is in accordance with the presumed left and right hemispheric divergence of metabolism and neuronal activation in depression [83, 84] suggesting that our results might have shown clearer trends in the TBS group in a larger sample [85].

Regarding clinical data of our population, the HAM-D scores at baseline in both the verum and sham groups were somewhat lower than in other studies investigating TMS in TRD [54, 86, 87], but are still reflective of an at least moderate depressive episode. The response rates were 25% in the verum group (2 out of 8), and 33.3% in the sham group (1 out of 3). While these rates seem to differ from recent studies by Berlim et al. and Voigt et al. [19, 88], a meta-analysis of Lepping et al. also finds high sham response rates [89]. The technology of transcranial magnetic stimulation undergoes constant refinement, hence studies on effects of TMS show great heterogeneity in treatment protocols (and duration) and inclusion criteria. Of note, in the current study recruitment was limited to patients currently not receiving treatment with mirtazapine, trazodone, quetiapine, aripiprazole, compatible for PET imaging of the 5-HT_1A_ receptor, as well as patients not receiving antiepileptic drugs or benzodiazepines on a regular basis regarding TBS treatment. Also, trajectories of remission and response to TMS seem to depend on specific characteristics, including age, benzodiazepine use and baseline depression severity [90]. An adequate level of functioning represents one condition for TBS, particularly in an outpatient setting, allowing for the inclusion of less severely depressed TRD patients (in comparison to, for example electroconvulsive therapy [91]). In addition, daily sessions might provide for a certain level of activation that could influence depression scores and symptom improvement over the treatment course. While treatment protocols for TBS are continuously refined based on new evidence, the herein reported response rates must be considered with caution, especially with our comparatively small sham group.

The size of the sham group (comprising female subjects only), but also the general sample size of 11 subjects constitutes the most important limitation of this study, restricting the generalizability of our data [92]. To increase statistical sensitivity, we focused our analysis on the bilateral DLPFCs where TBS was administered. However, other ROIs, especially the raphe, but also the hippocampus and amygdala, where changes in 5-HT_1A_ receptor might be expected, are missing in this analysis. Also, we did not account for potential effects of concomitant antidepressant pharmacotherapy in our analyses [93]. Given that treatment regimens had to remain unchanged throughout study participation, we expect these to be negligible.

In conclusion, we could show an effect of three-week bilateral TBS treatment on the distribution volumes of the 5-HT_1A_ receptor in a group of patients suffering from treatment resistant depression, with an increase in 5-HT_1A_ receptor availability correlating with a greater difference in depression score post-treatment. While these results appear indicative for a connection of the 5-HT_1A_ receptor with the mechanisms of action of theta-burst stimulation, they must be interpreted with caution, particularly because of the small sample and sham group size.

## Data Availability

Processed data are available from the corresponding author on reasonable request and a data-sharing agreement. Due to the data protection, raw data are not available

## ACKNOWLEDGEMENTS

This research was funded in whole, or in part, by the Austrian Science Fund (FWF) [Grant number KLI 551, PI: S. Kasper]. M. Murgaš is funded by the Austrian Science Fund (FWF) [Grant number DOC 33-B27, Supervisor R. Lanzenberger]. M.B. Reed is a recipient of a DOC fellowship of the Austrian Academy of Sciences at the Department of Psychiatry and Psychotherapy, Medical University of Vienna. We would like to express our gratitude towards Jonathan Downar for his assistance in the conceptualization of the study. We would further like to thank Gregor Gryglewski, Marius Hienert, Marie Spies, Christoph Kraus, Alexander Kautzky, Arkadiusz Komorowski, Paul Michenthaler, and Richard Frey for clinical support, and Lucas Rischka and Sebastian Ganger for technical support, and all additional staff and students from the Neuroimaging Lab (NIL) involved in the realization of this research. Moreover, we would like to thank radiotechnologists, Ingrid Leitinger and Harald Ibeschitz, for operating PET.

