## Supplementary material for "Effects of bilateral sequential theta-burst stimulation on 5-HT_1A_ receptors on dorsolateral prefrontal cortex in treatment resistant depression": Supplemetary Material

**Supplement**

| Subject | Diagnosis | Treatment group | Current Medication |
|---|---|---|---|
| 1 | Recurrent depressive disorder | Sham | Sertraline, Pregabalin |
| 2 | Recurrent depressive disorder | Bilateral TBS | Escitalopram, Pregabalin, Lithium, Quetiapine (up to 50mg as required) |
| 3 | Recurrent depressive disorder | Sham | Tranylcypromine, Duloxetine, Lithium, Lorazepam (up to 1,25mg as required) |
| 4 | Recurrent depressive disorder | Bilateral TBS | Tranylcypromine, Milnacipran, Melitracen, Flupentixol, Alprazolam (0.5mg as required) |
| 5 | Recurrent depressive disorder | Sham | Venlafaxine, Zolpidem, Lorazepam (up to 2mg daily as required) |
| 6 | Recurrent depressive disorder | Bilateral TBS | Sertraline |
| 7 | Major depressive episode | Bilateral TBS | Escitalopram, Mianserin |
| 8 | Recurrent depressive disorder | Bilateral TBS | Escitalopram |
| 9 | Recurrent depressive disorder, dysthymia | Bilateral TBS | Venlafaxine, Lithium |
| 10 | Major depressive episode | Bilateral TBS | Milnacipran |
| 11 | Recurrent depressive disorder | Bilateral TBS | Melitracen, Flupentixol |

*Table 1 Concomitant medication of study subjects, both depressive episode and recurrent depressive disorder diagnoses were therapy-resistant*
